# PRIMARY HEALTH CARE PERFORMANCE INDICATORS MODEL IN A PERFORMANCE-BASED CAPITATION SCHEME TO MEASURE NATIONAL HEALTH INSURANCE HEALTH SERVICE EQUITY (PROTOCOL STUDY)

**DOI:** 10.1101/2023.06.06.23291020

**Authors:** Ari Dwi Aryani, Adang Bachtiar, Cicilya Candi

**Affiliations:** Faculty of Public Health, University Indonesia, Depok, Indonesia

**Keywords:** Healthcare Equity, Healthcare Capacity, Primary Healthcare Performance

## Abstract

The inequity of health services still occurs after the implementation of the National Health Insurance (JKN). Regular monitoring of the Performance of Primary Health Care is the key to reducing the inequity of health services as the main goal of JKN. The application of Performance-Based Capitation (KBK) with three indicators since 2016, shows an improvement in Primary Health Care’s performance in improving quality and efficiency in the first level of service in comparison. Primary Health Care performance is influenced by Primary Health Care capacity and has an impact on the output of primary service performance (Health Service Equity). This study aims to develop a model of performance indicators, Primary Health Care capacity and equity indicators, in order to measure healthcare equity. The research design used an exploratory sequential-mixed method. The research is divided into three stages. Phase one is a systematic review to identify indicators that can be used in measuring the capacity, the performance of Primary Health Care and healthcare equity. Phase two is carried out with a qualitative approach with the Consensus Decision Making Group (CDMG) to determine indicators that can be used in measuring the capacity and performance of Primary Health Care as well as measuring health service equity with experts. Phase three is to develop a model of Primary Health Care performance indicators based on a capitation scheme that can measure the equity of healthcare access. This stage is carried out using Multilevel Structural Equation Modeling (MSEM) analysis. It is hoped that this model can be used by Social Security Administrator for Health (BPJS Kesehatan) and the Ministry of Health to measure the performance of Primary Health Care and health service equity.

## Introduction

Equity to access to healthcare is a challenge faced by many countries around the world. To achieve equitable access to health services, every country in the world is obliged to provide or bear the health insurance of its population in accordance with the main target of Sustainable Development Goals (SDGs) achieving Universal Health Coverage (UHC). This is in line with the implementation of the National Health Insurance (JKN) which is the state’s commitment to achieving UHC [1].

The principle of equity in the implementation of JKN according to the National Social Security System (SJSN) Law is the similarity of getting services according to medical needs regardless of how much contribution is paid. There has been an increase in access and utilization of health services since JKN began to be implemented in 2014. The utilization rate of health services reached 1.07 million per day, an increase of more than 100%. The utilization rate in Primary Health Care increased by 61.2 per cent to 140.47 per mile [2]. However, these achievements do not necessarily make JKN’s goal of achieving healthcare equity realized.

BPJS Kesehatan implements performance-based capitation payments (KBK), which aim to improve Primary Health Care performance through monitoring Primary Health Care performance and linking it to monthly capitation payments. Of the three Primary Health Care KBK performance indicators, only non-specialist referral ratio figures indicators were achieved nationally. Contact Number is an indicator to determine the level of accessibility and utilization of primary services in Primary Health Care by participants. But this indicator is assessed without taking into account geographical conditions and population density. So an indicator is needed that takes into account geographical conditions and the ratio of Primary Health Care compared to participants.

Another indicator is the Controlled Prolanis Participant Ratio, Prolanis is the abbreviation of ‘Program Pengelolaan Penyakit Kronis’ or Chronic Disease Management Program. This indicator is to measure the number of Prolanis DM and HT participants in Primary Health Care without taking into account the health condition of the participants and without strengthening promotional and preventive efforts. Currently, the preparation of new benefits of the JKN program is being carried out. The new benefit concept focuses on improving preventive efforts with increased health screening coverage. This concept is in line with the health transformation launched by the Ministry of Health. There was an increase in individual preventive services in the form of health screening for 14 catastrophic diseases. The new benefits in question are based on basic health needs with the aim of providing equitable and sustainable health services. So indicators are needed that can measure promotive and preventive efforts.

Within the framework of Primary Health Care performance monitoring, there are three interconnected domains, namely capacity, performance and output (quality, efficiency and equity). The performance of the first level of services is influenced by the capacity of the Primary Health Care and the output is the result of the performance of the Primary Health Care [3]. Several studies using a qualitative approach show that the performance achievement of KBK is influenced by the capacity of Primary Health Care which includes the availability of human resources, the availability of infrastructure, governance and organization, the availability of information systems, and financing [4]. In order to be able to carry out comprehensive performance monitoring, an Primary Health Care capacity indicator is needed.

The performance output in the KBK scheme shows an improvement in the quality of Primary Health Care, the quality index value achieved by Performance-based Primary Health Care is better than similar values in the Non-Performance-based Primary Health Care group. Although there is an increase in JKN’s first-level service performance nationally and has an impact on quality and efficiency, there are still differences in performance achievements between regions, per type of Primary Health Care and each Primary Health Care [1]. Therefore, it is necessary to assess the performance of Primary Health Care by region in order to measure equity.

This research is to develop a model of Primary Health Care performance indicators in the KBK scheme that can be used to periodically measure and monitor performance. So that in the end, it can increase the equity of health services.

**Research Question**

- What are the Primary Health Care capacity and performance indicators?
- What is the equity measurement indicator of Primary Health Care?
- What is the Primary Health Care performance indicator model in the KBK scheme that can measure healthcare equity?

## Materials and Methods

### Conceptual Framework

This research is expected to provide updates in the form of a KBK performance indicator model and the weight of assessing the calculation of performance achievements in accordance with the new JKN Program benefit policy based on basic health needs.

This conceptual framework is structured in reference to the theoretical framework which is a modification between the research primary service monitoring framework of Erica Barbazza (2019), Dionne Kringos (2020) and Ebert (2017). The primary service monitoring framework that is compiled consists of 3 dimensions, namely Primary Health Care capacity, Primary Health Care performance and primary service results (Fig.1).

**Figure 1.**
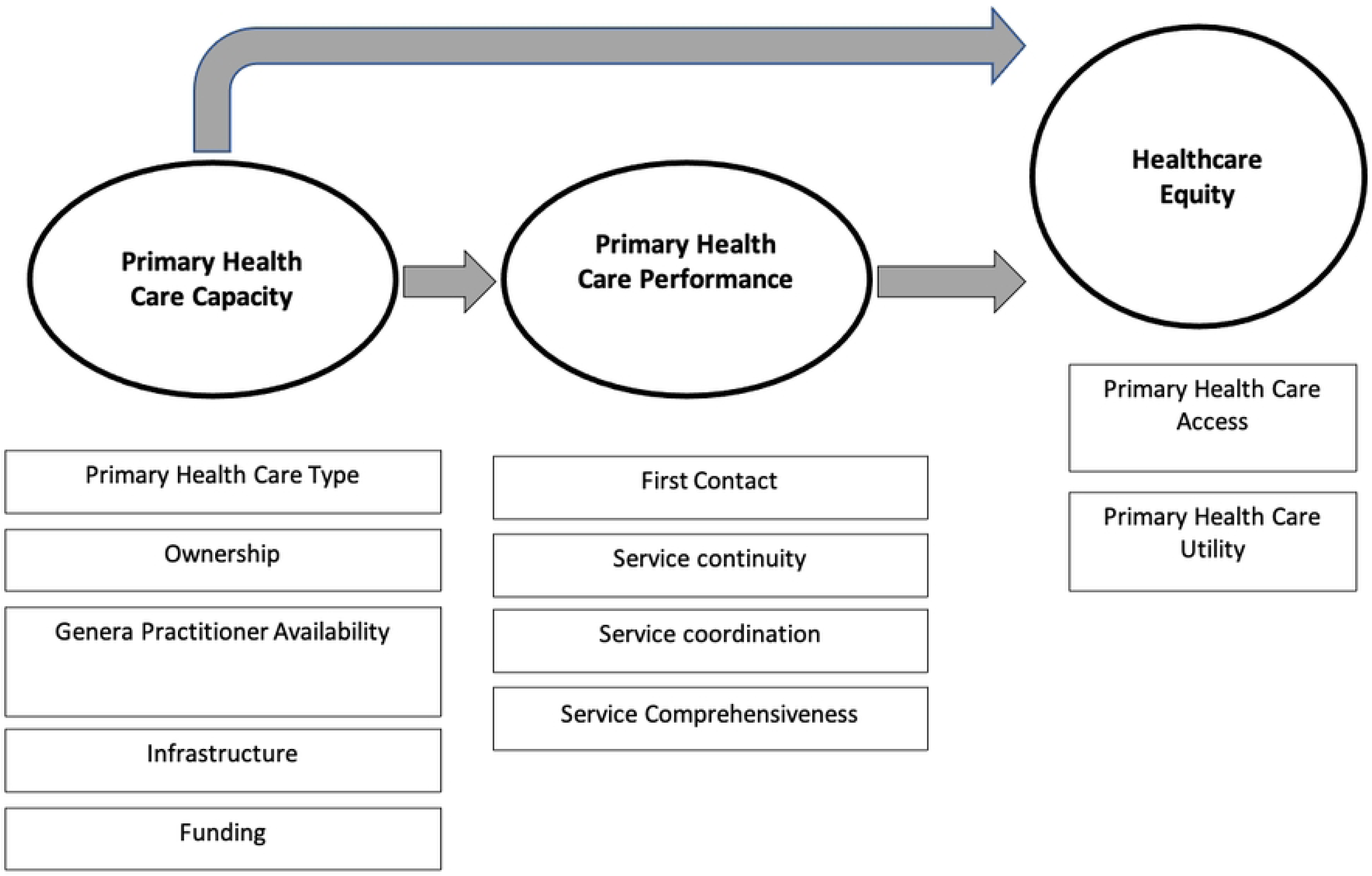
Conceptual Framework.

### Research Design

The research method used is mixed methods with qualitative methods and quantitative methods. There are several reasons to use mixed methods in health science research. Researchers can seek to look at problems from multiple perspectives to enhance and enrich the meaning of a single perspective. Researchers may also want to contextualise information, to take macro pictures of a system and additional information about individuals. Another reason to combine quantitative and qualitative data is to develop a complete understanding of a problem; develop complementary images; compare, validate, or triangulate results; provide context illustrations for trends; or to examine the process of shared experience (Plano Clark, 2010 in John W. Creswell, 2018). The research consists of 3 phases (Fig. 2) are described below (Table. 1). All the operational definitions are explained underneath (Table. 2).

**Table 1.** Description of study.

| Description | Phase of study |  |  |
| --- | --- | --- | --- |
|  | 1st | 2nd | 3rd |
| Output | Obtain capacity indicators, Primary Health Care performance indicators, health service equity measurement indicators and Primary Health Care performance measurement framework. | Determine Primary Health Care capacity and performance indicators, indicators to measure health service equity | Obtain and develop equity measurement models |
| Data Collection | *Systematic literature* | Consensus Decision | Test the Primary |
|  | *review* | Making Group (CDMG) | Health Care measurement framework and Primary Health Care KBK performance indicators. |
| Subject | Literature (2013-2022) from PubMed, Google Scholar, PubMed Central, Researchgate, Elsevier and other sources.<br>Keywords: indicator, performance, capacity, equity, primary care. | Health Ministry, DJSN, BPJS Kesehatan, Professional organizations, Primary healthcare Associations, Primary healthcare management and Academics | Secondary data taken from the national database of BPJS Kesehatan will be carried out in January 2023. |
| Sampling |  | Purposive sampling | Total sampling |
| Data References | Inclusion: Articles or journals published in Indonesian and English. Published from 2017 to 2022.<br>Exclusion: Studies are excluded if they are without the full text available and which are not written in Indonesian or English | Results of CDMG | Inclusion:<br>• All KBK achievement data for 2018-2020<br>• Data on primary outpatient services and referrals of JKN Participants during 2018 - 2020 from all Primary health care and hospitals in collaboration with BPJS Kesehatan nationally<br>• All types of Primary health care, ownership status, primary health care credential results,<br>capitation norms, and capitation payments.<br>Exclusion: CDMG results and according to the literature. |
| Analysis | Articles are selected based on PICO criteria. Furthermore, screening is carried out for the possibility of duplication of articles that have been searched. Screening is done based on the title and abstract. Then, the qualified articles are included in the qualitative synthesis. | Thematic analysis aims to present the model of intervention proposal with the CDMG approach with the subject of study. | Univariate, Multilevel Regression, Structural Equation Model (SEM) Partial Least Square (PLS), and Multilevel Structural Equation Model (MSEM) |

**Table 2.** Operational definitions and measurement scale.

| Variable | Definition | Measurement method suggestion | Measurement scale | Scale |
| --- | --- | --- | --- | --- |
| **Primary Health Care Performance** (based on functions and benefits) |  |  |  |  |
| First contact primary health care | The first visit of the patient when experiencing complaints/health problems [5] | Contact number<br>Source: BPJS Kesehatan Health Facilities Information System (HFIS) Database | (Will be determined on *Consensus Decision Making Group*) |  |
| Service continuity | Continuous services are provided to patients in an effort to prevent, treat, manage and control the disease [6] | 1. Controlled Prolanist Participant Ratio [7]<br>2. The proportion of diabetic patients with HbA1c levels of | (Will be determined on *Consensus Decision Making Group*) |  |
|  |  | <7.5% in the previous 12 months [8] [10] [9]<br>3. The proportion of Hypertensive patients with controlled blood pressure [8]<br>Source: BPJS Kesehatan Health Facilities Information System (HFIS) Database |  |  |
| Service coordination | Coordinated service delivery between Primary health care and Referral Health Facilities [6] | 1. % of patients who come without being referred to a hospital [8] [3]<br>2. Non-Specialist referral ratio [7]<br>3. % of Patients refer back [8]<br>Source: BPJS Kesehatan Database | (Will be determined on *Consensus Decision Making Group*) |  |
| Service Comprehensiveness | Provision of comprehensive health services to prevent, treat, manage and control disease [6] | 1. Pain rate of Participants Registered at Primary Health Care<br>2. Prevalence of Chronic Obstructive Pulmonary Disease Patients [11] [3]<br>3. Prevalence of Heart Disease Patients [11]<br>4. Prevalence of cancer patients [11]<br>5. Hypertension Treatment Coverage [3]<br>TB Treatment Coverage [3]<br>6. % Breast Cancer Screening [8] | (Will be determined on *Consensus Decision Making Group*) |  |
|  |  | 7.% Cervical Cancer Screening[8]<br>8.% Heart Disease Screening [12]<br>9.% DM Screening [12]<br>10. % Hypertension Screening [12]<br>Diabetes Education [3]<br>Source: BPJS Kesehatan Database |  |  |
| **Primary Health Care Capacity** |  |  |  |  |
| Primary Health Care Type | The type of Primary Health Care is based on the grouping determined by the Ministry of Health | Basis Data *Health Facilities Information System* (HFIS) BPJS Kesehatan | Primary Health Care based on type.<br>1 = Puskesmas<br>2 = Klinik Pratama<br>3 = RSD Pratama<br>4 = Dokter Praktik Perorangan | Nominal |
| Ownership | The assessment of Primary health care management is seen from its ownership | Basis Data *Health Facilities Information System* (HFIS) BPJS Kesehatan | Primary health care Ownership<br>1 = Government<br>2 = Private | Nominal |
| General Practitioner Availability | The availability of General Practitioners in Primary health care is measured based on the ratio of the number of Doctors compared to the number of participants served [5] | Basis Data *Health Facilities Information System* (HFIS) BPJS Kesehatan |  | Ratio |
| Infrastructure | The feasibility of health facilities is assessed | Basis Data *Health Facilities Information* | 1 = Score >70 Recommended | Ordinal |
|  | based on the number of Primary health care credentialing scores [5] | *System* (HFIS) BPJS Kesehatan | 2 = Skor <70<br>Not Recommended |  |
| Funding | The amount of capitation acceptance given is based on performance appraisal [5] | Basis Data *Health Facilities Information System* (HFIS) BPJS Kesehatan | 1 = acceptance of capitation 100%<br>2 = acceptance of capitation < 100% | Ordinal |
| Healthcare Equity | Similarities in access and utilization of health services between groups based on socio-economic level, age, sex [5] [6] |  |  |  |
| **Service Access (Vased on the place of health facilities)** |  |  |  |  |
| Primary healthcare Location | The location of Primary Health Care is in remote or non-remote areas | Basis Data *Health Facilities Information System* (HFIS) BPJS Kesehatan | 1 = Non Isolated<br>2 = Isolated | Nominal |
| Utilization Number of contacts per participant region | The number of people who avail the service in a given period of time | Number of contacts per region<br>BPJS Kesehatan Health Service Database | (Will be determined on *Consensus Decision Making Group*) |  |
| Utilization by Demographics | Number of people utilizing the service in a given time period based on demographics<br>1. Age<br>2. Gender<br>3. Social-Economy (PBI/Non-PBI)<br>4. Location (City/Town) | BPJS Kesehatan Health Service Database | (Will be determined on *Consensus Decision Making Group*) |  |

**Figure 2.**
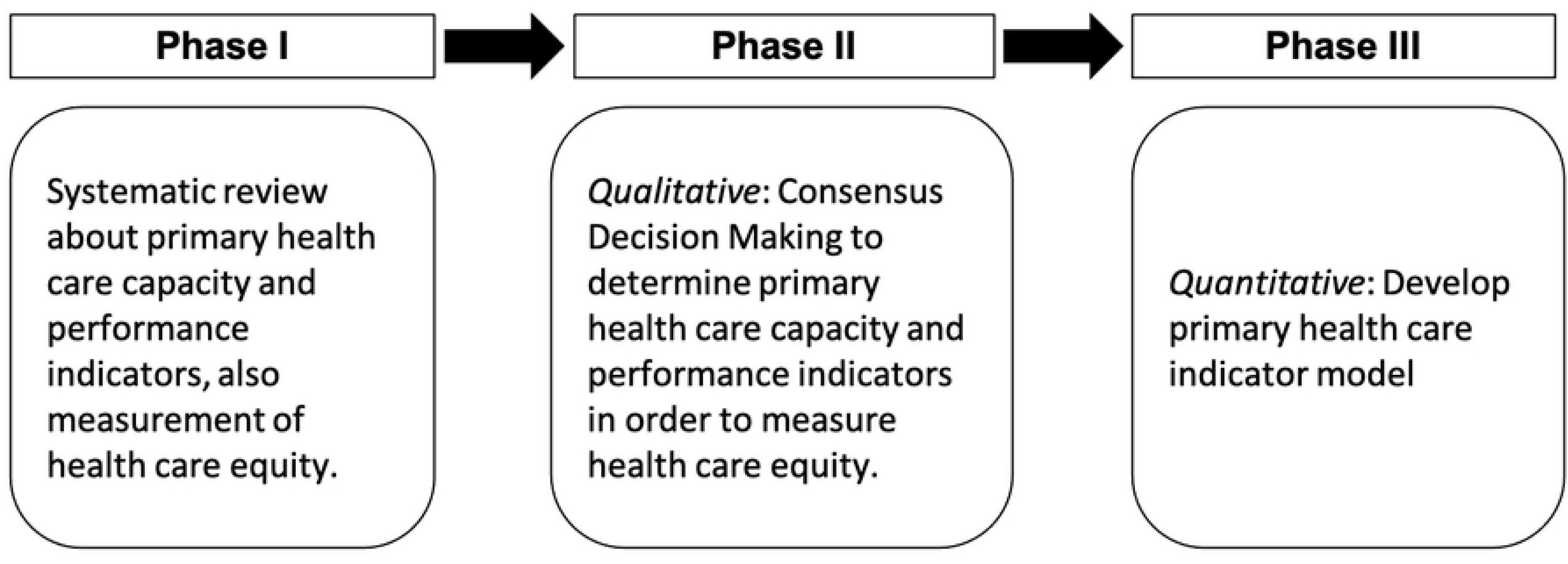
Phase of The Study.

### Ethical Research

This research has been approved by the Research and Community Service Ethics Committee of the Faculty of Public Health, University of Indonesia (no. Ket-31/UN2.F10.D11/PPM.00.02/2023) From Jan 2023 – Feb 2023

## Data Availability

No datasets were generated or analysed during the current study. All relevant data from this study will be made available upon study completion.

## Author contributions

**Conceptualization:** Ari Dwi Aryani, Adang Bachtiar

**Funding acquisition:** Ari Dwi Aryani

**Methodology:** Ari Dwi Aryani, Adang Bachtiar

**Writing (original draft)**: Ari Dwi Aryani

**Writing (review & editing)**: Adang Bachtiar, Cicilya Candi

